# Results of safety monitoring of BNT162b2 (Pfizer-BioNTech) COVID-19 vaccine in U.S. children aged 5-17 years

**DOI:** 10.1101/2022.10.28.22281532

**Authors:** Mao Hu, Hui Lee Wong, Yuhui Feng, Patricia C. Lloyd, Elizabeth R. Smith, Kandace L. Amend, Annemarie Kline, Daniel C. Beachler, Joann F. Gruber, Mahasweta Mitra, John D. Seeger, Charlalynn Harris, Alex Secora, Joyce Obidi, Jing Wang, Jennifer Song, Cheryl N. McMahill-Walraven, Christian Reich, Rowan McEvoy, Rose Do, Yoganand Chillarige, Robin Clifford, Danielle D Cooper, Azadeh Shoaibi, Richard Forshee, Steven A. Anderson

## Abstract

**Importance:** Active monitoring of health outcomes following COVID-19 vaccination offers early detection of rare outcomes that may not be identified in pre-licensure trials.

**Objective:** To conduct near-real time monitoring of health outcomes following BNT162b2 COVID-19 vaccination in the U.S. pediatric population aged 5-17 years.

**Design:** We conducted rapid cycle analysis of 20 pre-specified health outcomes, 13 of which underwent sequential testing and 7 of which were monitored descriptively within a cohort of vaccinated individuals. We tested for increased risk of each health outcome following vaccination compared to a historical baseline, while adjusting for repeated looks at the data as well as claims processing delay.

**Setting:** This is a population-based study in three large commercial claims databases conducted under the U.S. FDA public health surveillance mandate.

**Participants:** The study included over 3 million enrollees aged 5-17 years with BNT162b2 COVID-19 vaccination through mid-2022 in three commercial claims databases. We required continuous enrollment in a medical health insurance plan from the start of an outcome-specific clean window to the COVID-19 vaccination.

**Exposure:** Exposure was defined as receipt of a BNT162b2 COVID-19 vaccine dose. The primary analysis assessed primary series doses together (Dose 1 + Dose 2), and dose-specific secondary analyses were conducted. Follow up time was censored for death, disenrollment, end of risk window, end of study period, or a subsequent vaccine dose.

**Main Outcome(s) and Measure(s):** We monitored 20 pre-specified health outcomes. We performed descriptive monitoring for all outcomes and sequential testing for 13 outcomes.

**Results:** Among 13 health outcomes evaluated by sequential testing, 12 did not meet the threshold for a statistical signal in any of the three databases. In our primary analysis, myocarditis/pericarditis signaled following primary series vaccination with BNT162b2 in ages 12-17 years across all three databases.

**Conclusions and Relevance:** Consistent with published literature, our near-real time monitoring identified a signal for only myocarditis/pericarditis following BNT162b2 COVID-19 vaccination in children aged 12-17 years. This method is intended for early detection of safety signals. Our results are reassuring of the safety of the vaccine, and the potential benefits of vaccination outweigh the risks.

**Key Points:** *Question:* Did active monitoring detect potentially elevated risk of health outcomes following BNT162b2 COVID-19 vaccination in the U.S. pediatric population aged 5-17 years?

*Findings:* Twelve of 13 health outcomes did not meet the safety signal threshold following BNT162b2 COVID-19 vaccination in three large commercial claims databases using near real-time monitoring. Myocarditis/pericarditis met the statistical threshold for a signal following primary series vaccination in ages 12-17 years.

*Meaning:* Results from near-real time monitoring of health outcomes following BNT162b2 COVID-19 vaccination provide additional reassuring evidence of vaccine safety in the pediatric population. The myocarditis/pericarditis signal is consistent with current evidence and is being further evaluated.

## Introduction

Three vaccines are currently available in the United States to prevent Coronavirus Disease 2019 (COVID-19) in children including Pfizer-BioNTech (BNT162b2), Moderna (mRNA-1273) for ages 6 months - 17 years, and Novavax (NVX-CoV2373) COVID-19 Vaccine ages 12-17 years.^1-3^ We present safety results from near-real time monitoring using commercial claims databases of over 3 million children aged 5-17 years following administration of the COVID-19 BNT162b2 vaccine, which was the first approved vaccine for the pediatric population. This study was conducted under the U.S. Food and Drug Administration (FDA) Biologics Effectiveness and Safety (BEST) Initiative using a rapid cycle analysis or near real-time monitoring framework which enables early detection of potential safety signals. This near real-time surveillance effort is part of the US government’s larger ongoing post-authorization monitoring and real-world evidence to ensure the safety of COVID-19 vaccines in children.

## Methods

### Data Sources

This study used administrative claims data from Optum, HealthCore (HealthCore Integrated Research Database), and CVS Health (Aetna Enterprise Data Warehouse) containing longitudinal medical and pharmacy claims data supplemented with vaccination data from participating local and state Immunization Information Systems (IIS) (Supplemental Table 1).^4^

### Study Population

We included health plan members aged 5-17 years who received BNT162b2 vaccination from the earliest date of emergency use authorization by age group through 6/25/2022 (Optum), 5/6/2022 (HealthCore), and 5/31/2022 (CVS Health). The study population was divided into three age groups according to the vaccine authorization schedule (5-11, 12-15, and 16-17 years). We required continuous enrollment in a medical health insurance plan from the start of an outcome-specific clean window to the COVID-19 vaccination.

### Exposure and Follow Up

Exposure was defined as the receipt of BNT162b2 COVID-19 vaccine (Supplemental Table 2). Dose number was assigned chronologically. The primary analyses included primary series doses (Dose 1 + Dose 2). Secondary analyses included stratification by Dose 1, 2, 3/booster. Follow up began on the date of eligible vaccination and was censored at subsequent vaccination, death, disenrollment, end of risk window, or end of study period.

### Health Outcomes

Using claims-based algorithms, we monitored 20 health outcomes selected through clinical consultation and literature review. Among them, 13 were assessed using sequential testing and seven were monitored descriptively due to lack of historical comparator.^5^ Myocarditis/pericarditis was assessed with varying risk windows and care settings based on evidence from prior surveillance efforts and clinician input (Supplemental Table 3).

### Sequential Testing

We conducted monthly sequential testing using the Poisson Maximized Sequential Probability Ratio Test^6^ and generated incidence rate ratios (RR) of observed outcome rates compared to database-specific historical (expected) rates. Pre-COVID-19-vaccination historical rates were adjusted for claims processing delay and standardized by age and sex where case counts permitted.^7^ One-tailed tests were used with a null hypothesis that the observed rate was no greater than the historical comparator rate beyond a pre-specified test margin with an overall alpha of 1%. A statistical signal occurred if the log likelihood ratio exceeded a critical value. Surveillance continued until a signal was detected or the prespecified maximum surveillance length was reached. ^5^

### Medical Record Review

Medical record review was conducted for myocarditis/pericarditis to assess the validity of cases identified by claims-based algorithms. Clinical adjudicators used the Brighton collaboration definition to classify cases, and records meeting the confirmed or probable Brighton classifications were considered a true case.^8^

## Results

### Descriptive Monitoring

Approximately 5.9 million BNT162b2 COVID-19 vaccine doses were administered to over 3 million commercial health plan enrollees aged 5-17 years across the three databases; 1,999,550 doses were observed in 1,000,895 enrollees in CVS Health; 2,033,212 doses in 1,078,712 enrollees in HealthCore; and 1,869,063 doses in 937,745 enrollees in Optum database (Figure 1). Demographic characteristics of the vaccinated populations were largely similar across databases (Table 1). We observed low incidence for the seven outcomes monitored descriptively in all three databases (<25 events of each outcome in each database; Supplemental Table 5).

**Figure 1.**
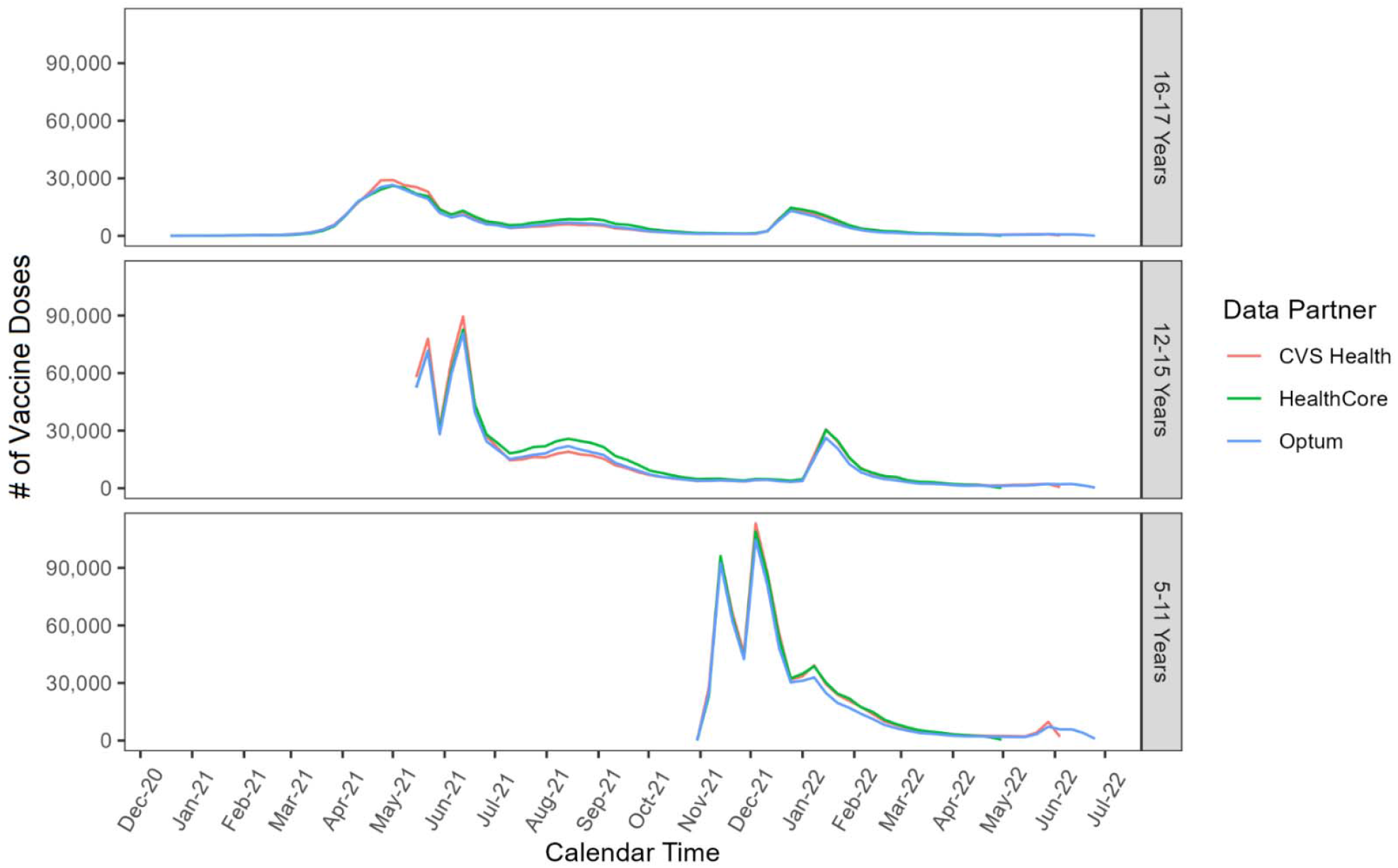
All Vaccine Doses Administered Among Recipients Aged 5-17 Years in CVS Health, HealthCore, and Optum Databases; FDA BEST System. *Study start date was the earliest EUA date for the BNT162b2 vaccination for each age group:*

- *Age 5-11: 10/29/2021*
- *Age 12-15: 5/10/2021*
- *Age 16-17: 12/11/2020* *Data cuts: CVS Health data through 5/31/2022, HealthCore data through 5/6/2022, and Optum data through 6/25/2022*

**Table 1.**
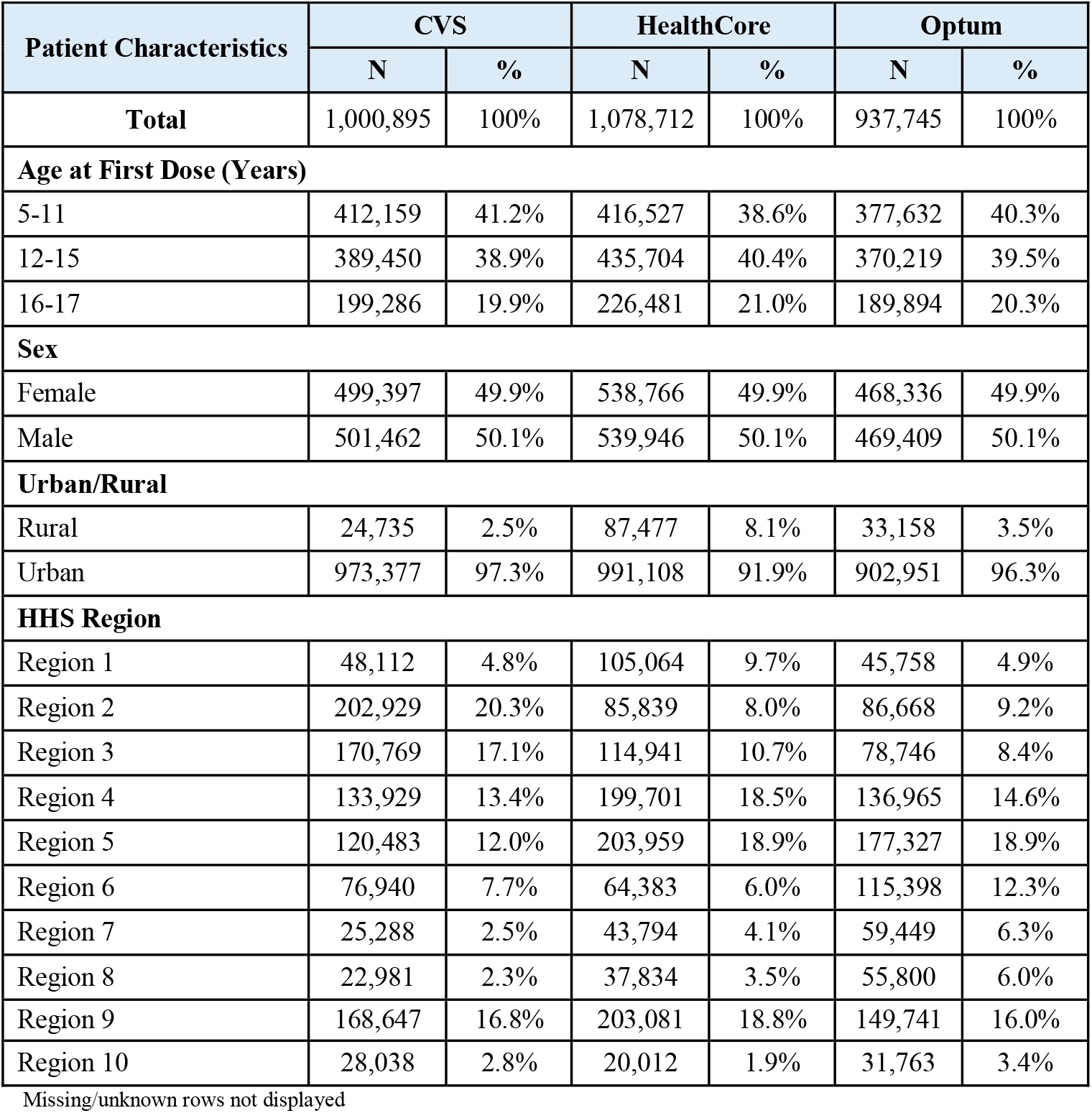
Characteristics of Health Plan Members Aged 5-17 Receiving BNT162b2 COVID-19 Vaccine in CVS Health, HealthCore, and Optum Databases.

| Patient Characteristics | CVS |  | HealthCore |  | Optum |  |
| --- | --- | --- | --- | --- | --- | --- |
|  | N | % | N | % | N | % |
| **Total** | 1,000,895 | 100% | 1,078,712 | 100% | 937,745 | 100% |
| **Age at First Dose (Years)** |  |  |  |  |  |  |
| 5-11 | 412,159 | 41.2% | 416,527 | 38.6% | 377,632 | 40.3% |
| 12-15 | 389,450 | 38.9% | 435,704 | 40.4% | 370,219 | 39.5% |
| 16-17 | 199,286 | 19.9% | 226,481 | 21.0% | 189,894 | 20.3% |
| **Sex** |  |  |  |  |  |  |
| Female | 499,397 | 49.9% | 538,766 | 49.9% | 468,336 | 49.9% |
| Male | 501,462 | 50.1% | 539,946 | 50.1% | 469,409 | 50.1% |
| **Urban/Rural** |  |  |  |  |  |  |
| Rural | 24,735 | 2.5% | 87,477 | 8.1% | 33,158 | 3.5% |
| Urban | 973,377 | 97.3% | 991,108 | 91.9% | 902,951 | 96.3% |
| **HHS Region** |  |  |  |  |  |  |
| Region 1 | 48,112 | 4.8% | 105,064 | 9.7% | 45,758 | 4.9% |
| Region 2 | 202,929 | 20.3% | 85,839 | 8.0% | 86,668 | 9.2% |
| Region 3 | 170,769 | 17.1% | 114,941 | 10.7% | 78,746 | 8.4% |
| Region 4 | 133,929 | 13.4% | 199,701 | 18.5% | 136,965 | 14.6% |
| Region 5 | 120,483 | 12.0% | 203,959 | 18.9% | 177,327 | 18.9% |
| Region 6 | 76,940 | 7.7% | 64,383 | 6.0% | 115,398 | 12.3% |
| Region 7 | 25,288 | 2.5% | 43,794 | 4.1% | 59,449 | 6.3% |
| Region 8 | 22,981 | 2.3% | 37,834 | 3.5% | 55,800 | 6.0% |
| Region 9 | 168,647 | 16.8% | 203,081 | 18.8% | 149,741 | 16.0% |
| Region 10 | 28,038 | 2.8% | 20,012 | 1.9% | 31,763 | 3.4% |

Missing/unknown rows not displayed

### Sequential Testing

Of the 13 outcomes sequentially tested, only myocarditis/pericarditis met the threshold for a statistical signal in any of the three databases. In the primary series analyses, myocarditis/pericarditis signaled in all databases for ages 12-15 and 16-17 years following BNT162b2 COVID-19 vaccination in all definitions of the myocarditis/pericarditis outcome (Table 2). In the dose-specific analyses, a signal was detected following Dose 2 in all definitions of the myocarditis/pericarditis outcome in ages 12-15 and 16-17 years in all data partners.

**Table 2.**
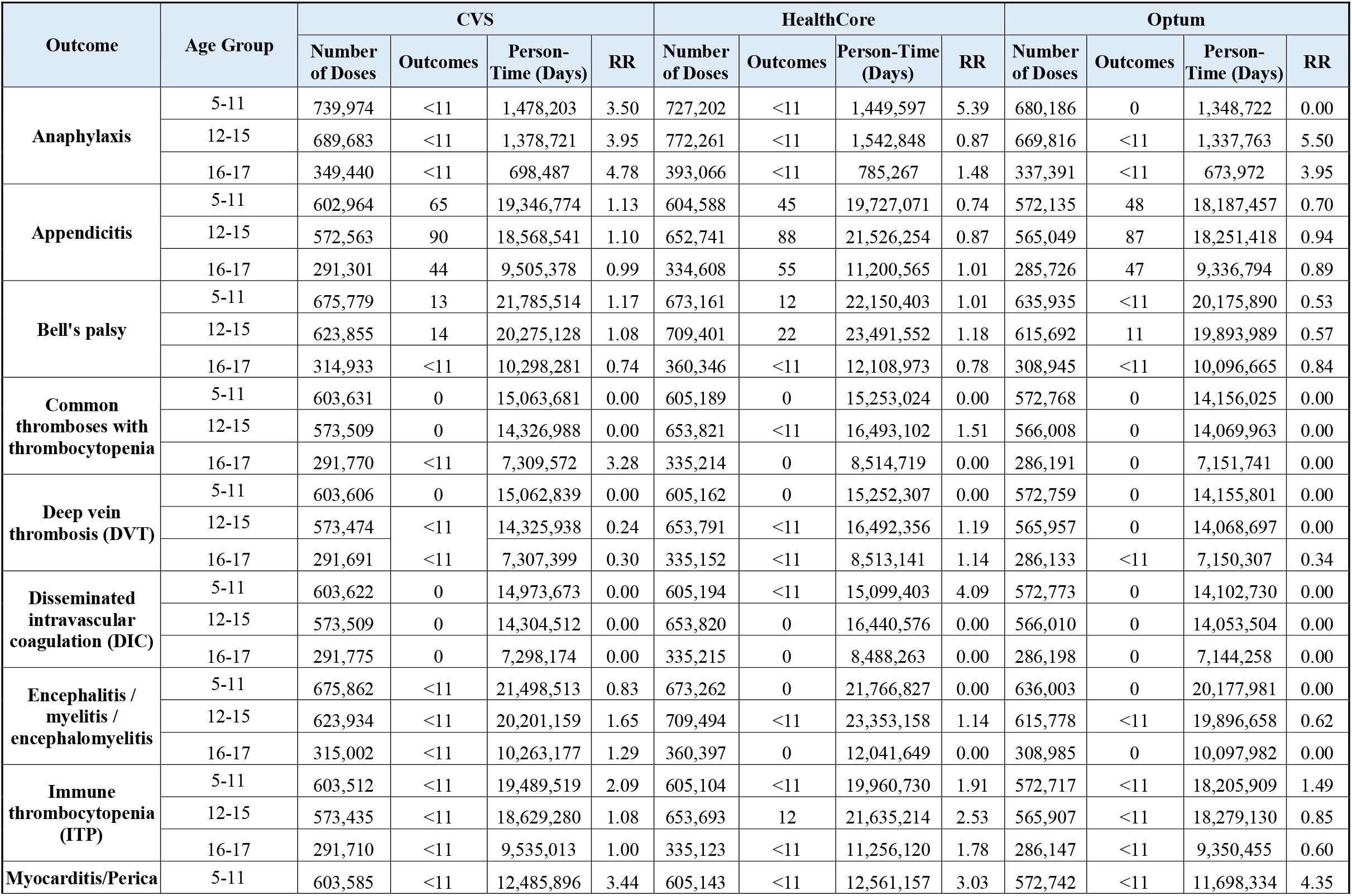
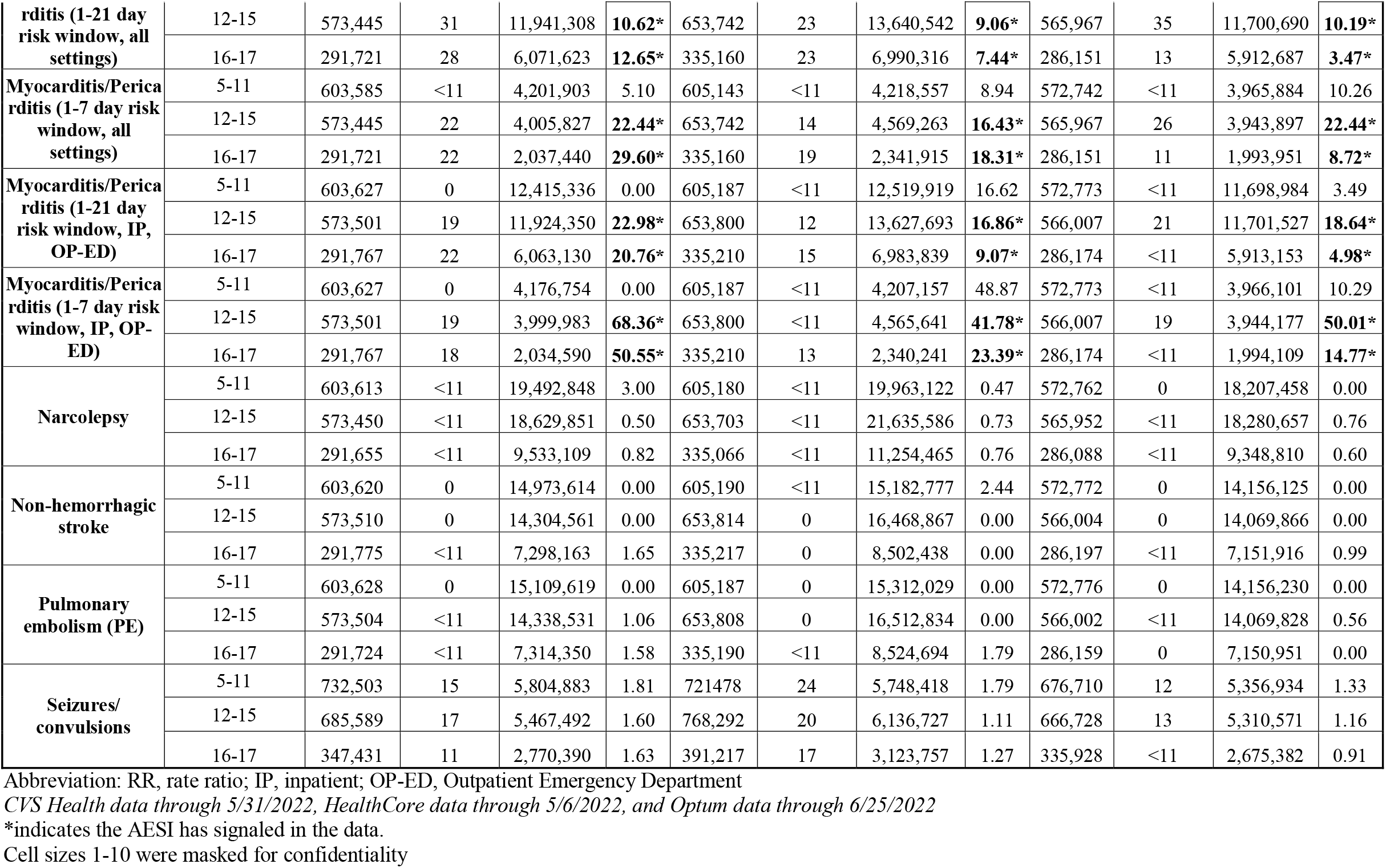
Sequential Testing Results in Health Plan Members Aged 5-17 years by AESI Following BNT162b2 Primary Series Doses in CVS Health, HealthCore, and Optum Databases.

Following Dose 3, signals were detected for some of the definitions of the myocarditis/pericarditis outcome in ages 16-17 years in CVS Health and ages 12-15 years in HealthCore databases (Supplemental Table 4). No signals were observed in ages 5-11.

### Medical Record Review

Of the 153 identified cases of myocarditis/pericarditis aged 12-17, medical record review was conducted for a sample of 37 cases whose charts were obtainable. Twenty-seven cases (73.0%) were confirmed as true myocarditis/pericarditis cases, of which 23 were male (93%) and 19 (70.4%) were hospitalized with a mean length of hospital stay of 2.8 days (median = 2 days). The mean time from vaccination to presentation for care for myocarditis/pericarditis was 6.8 days (median = 3 days).

## Discussion

Our near real-time monitoring results of 20 pre-specified health outcomes in the pediatric population provide reassuring real-world evidence of the safety of the BNT162b2 COVID-19 vaccine in children and adolescents. The signal detected for myocarditis/pericarditis is consistent with published peer-reviewed publications demonstrating an elevated risk of myocarditis/pericarditis following mRNA vaccines, especially among younger males aged 12-29 years.^9-11^ It should be noted that myocarditis/pericarditis is a rare event, with an average incidence of 39.4 cases per million doses administered in ages 5-17 years within 7 days after BNT162b2 COVID-19 vaccination.^12-13^ We did not detect a signal for myocarditis/pericarditis in young children aged 5-11 years, which is consistent with reports from other surveillance systems.^14-15^

This study has several strengths. First, the study included a large, geographically diverse population from three U.S. commercial health insurance databases. Due to availability of more complete information from claims supplemented with IIS data and a short data lag from health encounters, we were able to monitor BNT162b2 COVID-19 vaccine safety in a near real-time manner. Additionally, identified myocarditis/pericarditis cases were validated through medical record review.

The study also has some limitations. First, this study only covers data on a commercially insured pediatric population and may not be nationally representative. We used a rapid monitoring method designed for early detection of potentially increased risk of health outcomes with limited confounding adjustment. Therefore, results of this study do not establish a causal relationship between the vaccine and health outcomes; signals should be further evaluated. Furthermore, the study may have limited power to detect small increases in risk of outcomes in certain subgroups with more recent authorizations such as booster doses in younger children. Analyses were not stratified by sex, which is an important demographic characteristic for certain outcomes.

FDA continues to monitor the safety of COVID-19 vaccines in the pediatric population and has expanded the framework to include additional age groups and vaccine brands with updated authorizations. The FDA BEST Initiative plays a major role in the larger U.S. federal government vaccine safety monitoring efforts and further supports decision-making concerning the safety of COVID-19 vaccines by healthcare professionals and the American public.

## Data Availability

The study protocol was publicly posted as referenced in the manuscript before data analyses and related documents can be made available where needed, by contacting the corresponding author. De-identified
participant data will not be shared without approval from the data partners.

## Additional Contributions

We thank Tainya C. Clarke and Kristin A. Sepúlveda of the US Food and Drug Administration; Anchi Lo, Bing Lyu, Derek Phan, Gyanada Acharya, Laurie Feinberg, Lloyd Marks, Minisha Kochar, Sandia Akhtar, Shruti Parulekar, Stella Zhu, William (Trey) Minter, Yeerae Kim, Yixin Jiao, and Zhiruo (Cassie) Wan of Acumen; Carla Brennan of CVS Health; Shiva Vojjala, Ramya Avula, Shiva Chaudhary, Shanthi P Sagare, Ramin Riahi, Navyatha Namburu, and Grace Stockbower of HealthCore; Michael Goodman, Michael Bruhn, and Ruth Weed of IQVIA; Grace Yang, Karen Schneider, Rebecca Braun, Megan Ketchell, and Kate Federici of Optum.

