## Supplementary Materials for "Results of safety monitoring of BNT162b2 (Pfizer-BioNTech) COVID-19 vaccine in U.S. children aged 5-17 years"

**Supplement (Online-only material)**

### Supplementary Table 1. Database Descriptions

| Database | Description | Claims Type | Update frequency | Data Lag,  Time to 80% Completeness^a^ | Enrollees  Ages 5-17 years^b^ | Count of IIS Jurisdictions Incorporated into Analysis |
| --- | --- | --- | --- | --- | --- | --- |
| CVS Health | CVS Health Clinical Trial Services (CVS CTS) transforms enrollment, demographic, and medical and drug claims data, for individuals enrolled from January 2018 forward in Aetna commercial including Affordable Care act (ACA) Marketplace, and Medicare Advantage health plans into a patient-centered, comprehensive Common Data Model (CDM). | Fully Adjudicated | Monthly | ~ 3-4 months for IP claims, 2-3 months for OP claims, and 1-2 months for professional claims | 5-11 years: > 1.5 million  12-15 years: > 991k  16-17 years: > 558k | 14 |
| Optum Pre-adjudicated Claims | The Optum data includes enrollment, prescription drug and pre-adjudicated hospital and physician health insurance claims. The pre-adjudicated claims database includes claims for privately insured and Medicare Advantage enrollees. Hospital and physician claims undergo initial processing on a daily basis from a large number of providers across the US who accept patients with health insurance. | Pre-Adjudicated | Bi-Weekly | ~ 1-2 months for IP, OP, and professional claims | 5-11 years: > 1.3 million  12-15 years: > 840k  16-17 years: > 429k | 18 |
| HealthCore | HealthCore, Inc. is a wholly-owned, research subsidiary of Elevance Health, Inc., a holding company owning several large US health plans associated with Anthem Blue Cross Blue Shield. HealthCore has a US population database including individually insured by commercial and Medicare Advantage plans, the HealthCore Integrated Research Environment (HIRE), with longitudinal data on health plan enrollees. | Fully Adjudicated | Monthly | ~ 2-3 months for IP claims and 1-2 months for OP and professional claims | 5-11 years: > 1.8 million  12-15 years: > 1.2 million  16-17 years: > 647k | 0 |

*^a^ Data lag based on 2020 claims delay distribution*

*^b^ Average number of annual enrollees in a given age category between 2018-2020*

### Supplementary Table 2. Codes for COVID-19 Vaccine Administrations, Utilized in Claims and IIS data

| HCPCS/CPT Code | CVX Codes (IIS-Specific) | Manufacturer | Name | Age Group | Vaccine Administration Code | NDC 11 Labeler Product ID (Vial) | Dosing Interval |
| --- | --- | --- | --- | --- | --- | --- | --- |
| NA | NA | Pfizer | Pfizer-BioNTech COVID-19 Vaccine | 16+ years | NA | 00069-2025-01 00069-2025-10 00069-2025-25 | NA |
| 91308 | 219 | Pfizer | Pfizer-BioNTech COVID-19 Vaccine | 6 month - 5 years | 0081A (1st dose) | 59267-0078-01 59267-0078-02 59267-0078-04 | -21+ days between dose 1 and dose 2 |
|  |  |  |  |  | 0082A (second dose) |  |  |
| 91307 | 218 | Pfizer | Pfizer-BioNTech COVID-19 Pediatric Vaccine | 5-11 years | 0071A (1st dose) | 59267-1055-01 59267-1055-02 59267-1055-04 | -21+ days between dose 1 and dose 2 -For immunocompromised, 21+ days between dose 1 and dose 2, and 28+ days between dose 2 and additional primary dose (dose 3) |
|  |  |  |  |  | 0072A (2nd dose) |  |  |
|  |  |  |  |  | 0073A (3rd dose) |  |  |
|  |  |  |  |  | 0074A (booster dose) |  |  |
| 91305 | 217 | Pfizer | Pfizer-BioNTech COVID-19 Vaccine | 12+ years | 0051A (1^st^ dose) | 59267-1025-01 59267-1025-02 59267-1025-03 59267-1025-04 | -21+ days between dose 1 and dose 2 and 5+ months between dose 2 and third/booster dose - For immunocompromised, 21+ days between dose 1 and dose 2, 28+ days between dose 2 and dose 3. Additionally, booster dose recommended 3+ months after primary series |
|  |  |  |  |  | 0052A (2^nd^ dose) |  |  |
|  |  |  |  |  | 0053A (3^rd^ dose) |  |  |
|  |  |  |  |  | 0054A (booster dose) |  |  |
| 91300 | 208 | Pfizer | Pfizer-BioNTech COVID-19 Vaccine | 12+ years | 0001A (1^st^ dose) | 59267-1000-01 59267-1000-02 59267-1000-03 | -21+ days between dose 1 and dose 2 and 5+ months between dose 2 and third/booster dose - For immunocompromised, 21+ days between dose 1 and dose 2, 28+ days between dose 2 and dose 3. Additionally, booster dose recommended 3+ months after primary series |
|  |  |  |  |  | 0002A (2^nd^ dose) |  |  |
|  |  |  |  |  | 0003A (3^rd^ dose) |  |  |
|  |  |  |  |  | 0004A (booster dose) |  |  |

**Supplementary** **Table 3. Outcomes, Age Groups, Settings, Clean Windows, Risk Windows, and Analysis Type for the Pediatric Population**

| **Outcome** | **Age Group of Interest** | **Setting** | **Clean Window** | **Risk Window** | **Analysis Type** |
| --- | --- | --- | --- | --- | --- |
| **Pediatric Outcomes** | | | | | |
| Myocarditis/  Pericarditis | Ages 5-17 years | IP, OP/PB | 365 days | 1-7 days^[1]^ | Descriptive and Sequential Testing |
|  | Ages 5-17 years | IP, OP/PB | 365 days | 1-21 days^[2]^ | Descriptive and Sequential Testing |
|  | Ages 5-17 years | IP, OP-ED | 365 days | 1-7 days^[1]^ | Descriptive and Sequential Testing |
|  | Ages 5-17 years | IP, OP-ED | 365 days | 1-21 days^[2]^ | Descriptive and Sequential Testing |
| Guillain-Barré syndrome (GBS) | Ages 5-17 years | IP- primary position only | 365 days | 1-42 days^[3,4]^ | Descriptive Only |
| Multisystem inflammatory syndrome in children (MIS-C) | Ages 5-17 years | IP, OP-ED | 365 days | 1-42 days^[5]^ | Descriptive Only |
| Encephalitis / myelitis / encephalomyelitis | Ages 5-17 years | IP | 183 days | 1-42 days^[6]^ | Descriptive and Sequential Testing |
| Transverse myelitis | Ages 5-17 years | IP, OP-ED | 365 days | 1-42 days^[7]^ | Descriptive Only |
| Anaphylaxis | Ages 5-17 years | IP, OP-ED | 30 days | 0-1 day^[8,9]^ | Descriptive and Sequential Testing |
| Common thromboses with thrombocytopenia | Ages 5-17 years | [Definition below]** | 365 days | 1-28 days^[10]^ | Descriptive and Sequential Testing |
| Unusual site thrombosis (broad)with thrombocytopenia- cerebral and abdominal thrombosis | Ages 5-17 years | [Definition below]** | 365 days | 1-28 days^[11]^ | Descriptive Only |
| Seizures/Convulsions | Ages 5-17 years | IP, OP-ED | 42 days | 0-7 days^[12]^ | Descriptive and Sequential Testing |
| Bell’s palsy | Ages 5-17 years | IP, OP/PB | 183 days | 1-42 days^[13]^ | Descriptive and Sequential Testing |
| Deep vein thrombosis (DVT) | Ages 5-17 years | IP, OP/PB | 365 days | 1-28 days^[14-16]^ | Descriptive and Sequential Testing |
| Pulmonary embolism (PE) | Ages 5-17 years | IP, OP/PB | 365 days | 1-28 days^[14-16]^ | Descriptive and Sequential Testing |
| Disseminated intravascular coagulation (DIC) | Ages 5-17 years | IP, OP-ED | 365 days | 1-28 days^[17]^ | Descriptive and Sequential Testing |
| Immune thrombocytopenia (ITP) | Ages 5-17 years | IP, OP/PB | 365 days | 1-42 days^[18,19]^ | Descriptive and Sequential Testing |
| Kawasaki disease | Ages 5-17 years | IP, OP/PB | 365 days | 1-28 days^[20,21]^ | Descriptive Only |
| Narcolepsy | Ages 5-17 years | IP, OP/PB | 365 days | 1-42 days^[22-24]^ | Descriptive and Sequential Testing |
| Appendicitis | Ages 5-17 years | IP, OP-ED | 365 days | 1-42 days^[25,26]^ | Descriptive and Sequential Testing |
| Non-hemorrhagic stroke | Ages 5-17 years | IP | 365 days | 1-28 days^[27,28]^ | Descriptive and Sequential Testing |
| Hemorrhagic stroke | Ages 5-17 years | IP | 365 days | 1-28 days^[27,28]^ | Descriptive Only |
| Acute myocardial infarction | Ages 5-17 years | IP | 365 days | 1-28 days^[27,28]^ | Descriptive Only |

*Definitions: Clean Window is defined as an interval used to define incident outcomes where an individual enters the study cohort only if the outcome of interest did not occur during that interval. Risk Window is defined as an interval during which occurrence of the outcome of interest will be included in the analyses.*

*Setting Definitions: IP refers to inpatient facility claims. OP-ED refers to a subset of outpatient facility claims occurring in the emergency department. OP/PB refers to all outpatient facility claims, and professional/provider claims except those professional/provider claims with a laboratory place of service. For myocarditis/pericarditis, analyses were performed using two different risk windows (1-7 days; 1-21 days) and two different care settings (inpatient, outpatient and provider services; inpatient and outpatient-emergency department) based on clinician input and available literature*

**All outcomes were identified using ICD-10-CM Diagnosis codes.^29,30^ The classification of outcomes into those to be monitored descriptively and those monitored via sequential testing is based on the availability of estimable background rates for the outcomes and the expected frequency of events*

*** Both Common thromboses with thrombocytopenia and Unusual site thrombosis (broad) with thrombocytopenia are combined outcomes consisting of a thrombotic event (made up of other events such as acute myocardial infarction, deep vein thrombosis etc.,) and a thrombocytopenia event (defined in the IP, OP/PB setting). The overall setting definition for each outcome depends on individual setting definitions for each of these components*

### Supplementary Table 4. Sequential Testing Results in Health Plan Members Aged 5-17 years by Outcome Following BNT162b2 All Doses (Primary Series, Dose 1, Dose 2, and Third/Booster Dose) in CVS Health, HealthCore, and Optum Databases

| **Outcome** | **Dose** | **Age Group** | **CVS** | | | | **HealthCore** | | | | **Optum** | | | |
| --- | --- | --- | --- | --- | --- | --- | --- | --- | --- | --- | --- | --- | --- | --- |
|  |  |  | **# of Doses** | **Outcomes** | **Person-Time (Days)** | **RR** | **# of Doses** | **Outcomes** | **Person-Time (Days)** | **RR** | **# of Doses** | **Outcomes** | **Person-Time (Days)** | **RR** |
| Anaphylaxis | Primary Series | 5-11 | 739,974 | <11 | 1,478,203 | 3.50 | 727,202 | <11 | 1,449,597 | 5.39 | 680,186 | 0 | 1,348,722 | 0.00 |
|  |  | 12-15 | 689,683 | <11 | 1,378,721 | 3.95 | 772,261 | <11 | 1,542,848 | 0.87 | 669,816 | <11 | 1,337,763 | 5.50 |
|  |  | 16-17 | 349,440 | <11 | 698,487 | 4.78 | 393,066 | <11 | 785,267 | 1.48 | 337,391 | <11 | 673,972 | 3.95 |
|  | Dose 1 | 5-11 | 391,692 | <11 | 782,495 | 3.30 | 404,716 | <11 | 807,094 | 4.81 | 371,307 | 0 | 735,473 | 0.00 |
|  |  | 12-15 | 365,895 | <11 | 731,399 | 2.48 | 425,137 | <11 | 849,202 | 1.58 | 360,549 | <11 | 719,866 | 7.67 |
|  |  | 16-17 | 187,361 | <11 | 374,491 | 8.92 | 221,794 | <11 | 443,014 | 2.63 | 185,157 | <11 | 369,815 | 7.20 |
|  | Dose 2 | 5-11 | 348,282 | <11 | 695,708 | 3.73 | 322,486 | <11 | 642,503 | 6.14 | 308,879 | 0 | 613,249 | 0.00 |
|  |  | 12-15 | 323,788 | <11 | 647,322 | 5.61 | 347,124 | 0 | 693,646 | 0.00 | 309,267 | <11 | 617,897 | 2.98 |
|  |  | 16-17 | 162,079 | 0 | 323,996 | 0.00 | 171,272 | 0 | 342,253 | 0.00 | 152,234 | 0 | 304,157 | 0.00 |
|  | Dose 3 | 5-11 | 13,611 | 0 | 23,369 | 0.00 | 1,481 | 0 | 2,929 | 0.00 | 21,450 | 0 | 32,023 | 0.00 |
|  |  | 12-15 | 96,975 | <11 | 192,972 | 10.17 | 66,312 | <11 | 131,608 | 11.11 | 86,087 | 0 | 170,125 | 0.00 |
|  |  | 16-17 | 66,162 | 0 | 131,909 | 0.00 | 45,560 | 0 | 90,677 | 0.00 | 55,445 | <11 | 110,204 | 12.24 |
| Appendicitis | Primary Series | 5-11 | 602,964 | 65 | 19,346,774 | 1.13 | 604,588 | 45 | 19,727,071 | 0.74 | 572,135 | 48 | 18,187,457 | 0.70 |
|  |  | 12-15 | 572,563 | 90 | 18,568,541 | 1.10 | 652,741 | 88 | 21,526,254 | 0.87 | 565,049 | 87 | 18,251,418 | 0.94 |
|  |  | 16-17 | 291,301 | 44 | 9,505,378 | 0.99 | 334,608 | 55 | 11,200,565 | 1.01 | 285,726 | 47 | 9,336,794 | 0.89 |
|  | Dose 1 | 5-11 | 318,219 | 27 | 7,902,725 | 1.13 | 335,279 | 21 | 8,916,128 | 0.74 | 310,350 | 25 | 7,865,129 | 0.84 |
|  |  | 12-15 | 301,730 | 29 | 7,378,691 | 0.89 | 356,294 | 42 | 9,243,397 | 0.97 | 302,196 | 41 | 7,529,320 | 1.07 |
|  |  | 16-17 | 155,214 | 20 | 3,875,060 | 1.10 | 186,911 | 27 | 5,072,915 | 1.10 | 155,615 | 17 | 4,028,348 | 0.75 |
|  | Dose 2 | 5-11 | 284,745 | 38 | 11,444,049 | 1.12 | 269,309 | 24 | 10,810,943 | 0.73 | 261,785 | 24 | 10,322,328 | 0.62 |
|  |  | 12-15 | 270,833 | 61 | 11,189,850 | 1.24 | 296,447 | 46 | 12,282,857 | 0.80 | 262,853 | 47 | 10,722,098 | 0.86 |
|  |  | 16-17 | 136,087 | 24 | 5,630,318 | 0.91 | 147,697 | 28 | 6,127,650 | 0.93 | 130,111 | 30 | 5,308,446 | 1.00 |
|  | Dose 3 | 5-11 | 11,438 | 0 | 124,510 | 0.00 | 1,339 | 0 | 45,888 | 0.00 | 19,033 | 0 | 369,675 | 0.00 |
|  |  | 12-15 | 86,036 | 11 | 3,355,000 | 0.83 | 63,891 | <11 | 2,482,199 | 0.74 | 79,497 | <11 | 3,073,489 | 0.55 |
|  |  | 16-17 | 57,772 | <11 | 2,289,307 | 0.81 | 43,066 | <11 | 1,697,120 | 0.83 | 50,525 | 15 | 1,951,045 | 1.43 |
| Bell's palsy | Primary Series | 5-11 | 675,779 | 13 | 21,785,514 | 1.17 | 673,161 | 12 | 22,150,403 | 1.01 | 635,935 | <11 | 20,175,890 | 0.53 |
|  |  | 12-15 | 623,855 | 14 | 20,275,128 | 1.08 | 709,401 | 22 | 23,491,552 | 1.18 | 615,692 | 11 | 19,893,989 | 0.57 |
|  |  | 16-17 | 314,933 | <11 | 10,298,281 | 0.74 | 360,346 | <11 | 12,108,973 | 0.78 | 308,945 | <11 | 10,096,665 | 0.84 |
|  | Dose 1 | 5-11 | 357,777 | <11 | 8,944,746 | 0.87 | 374,144 | <11 | 10,005,282 | 1.28 | 345,766 | <11 | 8,760,032 | 0.24 |
|  |  | 12-15 | 328,898 | <11 | 8,079,623 | 0.97 | 386,972 | <11 | 10,101,801 | 1.01 | 329,457 | <11 | 8,231,955 | 0.50 |
|  |  | 16-17 | 167,972 | <11 | 4,216,467 | 0.61 | 201,636 | <11 | 5,513,254 | 0.38 | 168,583 | <11 | 4,378,145 | 1.11 |
|  | Dose 2 | 5-11 | 318,002 | <11 | 12,840,768 | 1.38 | 299,017 | <11 | 12,145,121 | 0.78 | 290,169 | <11 | 11,415,858 | 0.75 |
|  |  | 12-15 | 294,957 | <11 | 12,195,505 | 1.28 | 322,429 | <11 | 13,389,751 | 1.32 | 286,235 | <11 | 11,662,034 | 0.62 |
|  |  | 16-17 | 146,961 | <11 | 6,081,814 | 1.05 | 158,710 | <11 | 6,595,719 | 1.11 | 140,362 | <11 | 5,718,520 | 0.64 |
|  | Dose 3 | 5-11 | 12,471 | 0 | 164,697 | 0.00 | 1,441 | 0 | 50,751 | 0.00 | 20,761 | 0 | 409,823 | 0.00 |
|  |  | 12-15 | 91,094 | <11 | 3,623,831 | 0.93 | 66,062 | <11 | 2,656,746 | 1.10 | 83,933 | <11 | 3,243,633 | 1.66 |
|  |  | 16-17 | 62,499 | <11 | 2,503,089 | 1.08 | 45,412 | <11 | 1,825,062 | 0.63 | 54,106 | <11 | 2,086,728 | 0.60 |
| Common thromboses with thrombocytopenia | Primary Series | 5-11 | 603,631 | 0 | 15,063,681 | 0.00 | 605,189 | 0 | 15,253,024 | 0.00 | 572,768 | 0 | 14,156,025 | 0.00 |
|  |  | 12-15 | 573,509 | 0 | 14,326,988 | 0.00 | 653,821 | <11 | 16,493,102 | 1.51 | 566,008 | 0 | 14,069,963 | 0.00 |
|  |  | 16-17 | 291,770 | <11 | 7,309,572 | 3.28 | 335,214 | 0 | 8,514,719 | 0.00 | 286,191 | 0 | 7,151,741 | 0.00 |
|  | Dose 1 | 5-11 | 318,571 | 0 | 7,281,081 | 0.00 | 335,610 | 0 | 7,872,732 | 0.00 | 310,689 | 0 | 7,096,078 | 0.00 |
|  |  | 12-15 | 302,226 | 0 | 6,810,180 | 0.00 | 356,881 | 0 | 8,241,243 | 0.00 | 302,702 | 0 | 6,848,840 | 0.00 |
|  |  | 16-17 | 155,466 | <11 | 3,530,834 | 3.40 | 187,251 | 0 | 4,401,614 | 0.00 | 155,870 | 0 | 3,578,004 | 0.00 |
|  | Dose 2 | 5-11 | 285,060 | 0 | 7,782,600 | 0.00 | 269,579 | 0 | 7,380,292 | 0.00 | 262,079 | 0 | 7,059,947 | 0.00 |
|  |  | 12-15 | 271,283 | 0 | 7,516,808 | 0.00 | 296,940 | <11 | 8,251,859 | 3.02 | 263,306 | 0 | 7,221,123 | 0.00 |
|  |  | 16-17 | 136,304 | <11 | 3,778,738 | 3.17 | 147,963 | 0 | 4,113,105 | 0.00 | 130,321 | 0 | 3,573,737 | 0.00 |
|  | Dose 3 | 5-11 | 11,446 | 0 | 89,279 | 0.00 | 1,341 | 0 | 33,982 | 0.00 | 19,048 | 0 | 291,412 | 0.00 |
|  |  | 12-15 | 86,169 | 0 | 2,293,118 | 0.00 | 63,983 | 0 | 1,724,614 | 0.00 | 79,622 | 0 | 2,092,120 | 0.00 |
|  |  | 16-17 | 57,851 | 0 | 1,553,697 | 0.00 | 43,144 | 0 | 1,163,968 | 0.00 | 50,629 | 0 | 1,327,042 | 0.00 |
| Deep vein thrombosis (DVT) | Primary Series | 5-11 | 603,606 | 0 | 15,062,839 | 0.00 | 605,162 | 0 | 15,252,307 | 0.00 | 572,759 | 0 | 14,155,801 | 0.00 |
|  |  | 12-15 | 573,474 | <11 | 14,325,938 | 0.24 | 653,791 | <11 | 16,492,356 | 1.19 | 565,957 | 0 | 14,068,697 | 0.00 |
|  |  | 16-17 | 291,691 | <11 | 7,307,399 | 0.30 | 335,152 | <11 | 8,513,141 | 1.14 | 286,133 | <11 | 7,150,307 | 0.34 |
|  | Dose 1 | 5-11 | 318,556 | 0 | 7,280,652 | 0.00 | 335,593 | 0 | 7,872,323 | 0.00 | 310,683 | 0 | 7,095,938 | 0.00 |
|  |  | 12-15 | 302,208 | 0 | 6,809,690 | 0.00 | 356,865 | <11 | 8,240,878 | 1.43 | 302,674 | 0 | 6,848,207 | 0.00 |
|  |  | 16-17 | 155,423 | 0 | 3,529,742 | 0.00 | 187,219 | <11 | 4,400,848 | 1.77 | 155,836 | <11 | 3,577,199 | 0.34 |
|  | Dose 2 | 5-11 | 285,050 | 0 | 7,782,187 | 0.00 | 269,569 | 0 | 7,379,984 | 0.00 | 262,076 | 0 | 7,059,863 | 0.00 |
|  |  | 12-15 | 271,266 | <11 | 7,516,248 | 0.45 | 296,926 | <11 | 8,251,478 | 0.95 | 263,283 | 0 | 7,220,490 | 0.00 |
|  |  | 16-17 | 136,268 | <11 | 3,777,657 | 0.58 | 147,933 | <11 | 4,112,293 | 0.47 | 130,297 | <11 | 3,573,108 | 0.34 |
|  | Dose 3 | 5-11 | 11,446 | 0 | 89,251 | 0.00 | 1,339 | 0 | 33,926 | 0.00 | 19,046 | 0 | 291,366 | 0.00 |
|  |  | 12-15 | 86,161 | 0 | 2,292,866 | 0.00 | 63,976 | 0 | 1,724,418 | 0.00 | 79,614 | <11 | 2,091,896 | 1.69 |
|  |  | 16-17 | 57,840 | 0 | 1,553,387 | 0.00 | 43,134 | 0 | 1,163,714 | 0.00 | 50,621 | 0 | 1,326,846 | 0.00 |
| Disseminated intravascular coagulation (DIC) | Primary Series | 5-11 | 603,622 | 0 | 14,973,673 | 0.00 | 605,194 | <11 | 15,099,403 | 4.09 | 572,773 | 0 | 14,102,730 | 0.00 |
|  |  | 12-15 | 573,509 | 0 | 14,304,512 | 0.00 | 653,820 | 0 | 16,440,576 | 0.00 | 566,010 | 0 | 14,053,504 | 0.00 |
|  |  | 16-17 | 291,775 | 0 | 7,298,174 | 0.00 | 335,215 | 0 | 8,488,263 | 0.00 | 286,198 | 0 | 7,144,258 | 0.00 |
|  | Dose 1 | 5-11 | 318,566 | 0 | 7,238,894 | 0.00 | 335,613 | 0 | 7,813,485 | 0.00 | 310,692 | 0 | 7,067,211 | 0.00 |
|  |  | 12-15 | 302,226 | 0 | 6,798,069 | 0.00 | 356,881 | 0 | 8,212,755 | 0.00 | 302,703 | 0 | 6,838,690 | 0.00 |
|  |  | 16-17 | 155,469 | 0 | 3,524,012 | 0.00 | 187,252 | 0 | 4,385,669 | 0.00 | 155,874 | 0 | 3,573,645 | 0.00 |
|  | Dose 2 | 5-11 | 285,056 | 0 | 7,734,779 | 0.00 | 269,581 | <11 | 7,285,918 | 8.81 | 262,081 | 0 | 7,035,519 | 0.00 |
|  |  | 12-15 | 271,283 | 0 | 7,506,443 | 0.00 | 296,939 | 0 | 8,227,821 | 0.00 | 263,307 | 0 | 7,214,814 | 0.00 |
|  |  | 16-17 | 136,306 | 0 | 3,774,162 | 0.00 | 147,963 | 0 | 4,102,594 | 0.00 | 130,324 | 0 | 3,570,613 | 0.00 |
|  | Dose 3 | 5-11 | 11,444 | 0 | 87,578 | 0.00 | 1,341 | 0 | 32,828 | 0.00 | 19,048 | 0 | 230,388 | 0.00 |
|  |  | 12-15 | 86,170 | 0 | 2,248,385 | 0.00 | 63,982 | 0 | 1,665,851 | 0.00 | 79,622 | <11 | 2,062,986 | 10.82 |
|  |  | 16-17 | 57,854 | 0 | 1,536,756 | 0.00 | 43,144 | 0 | 1,139,733 | 0.00 | 50,630 | 0 | 1,317,447 | 0.00 |
| Encephalitis / myelitis / encephalomyelitis | Primary Series | 5-11 | 675,862 | <11 | 21,498,513 | 0.83 | 673,262 | 0 | 21,766,827 | 0.00 | 636,003 | 0 | 20,177,981 | 0.00 |
|  |  | 12-15 | 623,934 | <11 | 20,201,159 | 1.65 | 709,494 | <11 | 23,353,158 | 1.14 | 615,778 | <11 | 19,896,658 | 0.62 |
|  |  | 16-17 | 315,002 | <11 | 10,263,177 | 1.29 | 360,397 | 0 | 12,041,649 | 0.00 | 308,985 | 0 | 10,097,982 | 0.00 |
|  | Dose 1 | 5-11 | 357,821 | 0 | 8,833,757 | 0.00 | 374,206 | 0 | 9,883,921 | 0.00 | 345,807 | 0 | 8,761,121 | 0.00 |
|  |  | 12-15 | 328,944 | 0 | 8,041,758 | 0.00 | 387,018 | 0 | 10,027,824 | 0.00 | 329,507 | <11 | 8,233,208 | 1.50 |
|  |  | 16-17 | 168,011 | 0 | 4,196,563 | 0.00 | 201,662 | 0 | 5,473,212 | 0.00 | 168,607 | 0 | 4,378,790 | 0.00 |
|  | Dose 2 | 5-11 | 318,041 | <11 | 12,664,756 | 1.44 | 299,056 | 0 | 11,882,906 | 0.00 | 290,196 | 0 | 11,416,860 | 0.00 |
|  |  | 12-15 | 294,990 | <11 | 12,159,401 | 2.75 | 322,476 | <11 | 13,325,334 | 1.99 | 286,271 | 0 | 11,663,450 | 0.00 |
|  |  | 16-17 | 146,991 | <11 | 6,066,614 | 2.18 | 158,735 | 0 | 6,568,437 | 0.00 | 140,378 | 0 | 5,719,192 | 0.00 |
|  | Dose 3 | 5-11 | 12,473 | 0 | 134,559 | 0.00 | 1,441 | 0 | 47,953 | 0.00 | 20,764 | 0 | 409,895 | 0.00 |
|  |  | 12-15 | 91,108 | 0 | 3,473,475 | 0.00 | 66,067 | 0 | 2,508,168 | 0.00 | 83,940 | 0 | 3,243,927 | 0.00 |
|  |  | 16-17 | 62,513 | 0 | 2,441,902 | 0.00 | 45,421 | 0 | 1,765,174 | 0.00 | 54,118 | 0 | 2,087,194 | 0.00 |
| Immune thrombocytopenia (ITP) | Primary Series | 5-11 | 603,512 | <11 | 19,489,519 | 2.09 | 605,104 | <11 | 19,960,730 | 1.91 | 572,717 | <11 | 18,205,909 | 1.49 |
|  |  | 12-15 | 573,435 | <11 | 18,629,280 | 1.08 | 653,693 | 12 | 21,635,214 | 2.53 | 565,907 | <11 | 18,279,130 | 0.85 |
|  |  | 16-17 | 291,710 | <11 | 9,535,013 | 1.00 | 335,123 | <11 | 11,256,120 | 1.78 | 286,147 | <11 | 9,350,455 | 0.60 |
|  | Dose 1 | 5-11 | 318,509 | <11 | 7,961,202 | 2.04 | 335,563 | <11 | 8,994,830 | 2.38 | 310,663 | <11 | 7,872,969 | 0.57 |
|  |  | 12-15 | 302,187 | <11 | 7,406,353 | 1.09 | 356,812 | <11 | 9,297,471 | 1.96 | 302,645 | <11 | 7,540,525 | 0.41 |
|  |  | 16-17 | 155,435 | <11 | 3,889,573 | 0.82 | 187,203 | <11 | 5,103,168 | 3.29 | 155,845 | <11 | 4,034,298 | 0.46 |
|  | Dose 2 | 5-11 | 285,003 | <11 | 11,528,317 | 2.13 | 269,541 | <11 | 10,965,900 | 1.51 | 262,054 | <11 | 10,332,940 | 2.20 |
|  |  | 12-15 | 271,248 | <11 | 11,222,927 | 1.07 | 296,881 | <11 | 12,337,743 | 2.95 | 263,262 | <11 | 10,738,605 | 1.16 |
|  |  | 16-17 | 136,275 | <11 | 5,645,440 | 1.12 | 147,920 | <11 | 6,152,952 | 0.54 | 130,302 | <11 | 5,316,157 | 0.70 |
|  | Dose 3 | 5-11 | 11,441 | 0 | 150,770 | 0.00 | 1,340 | 0 | 47,511 | 0.00 | 19,043 | 0 | 369,817 | 0.00 |
|  |  | 12-15 | 86,150 | 0 | 3,430,922 | 0.00 | 63,971 | 0 | 2,573,226 | 0.00 | 79,614 | 0 | 3,078,046 | 0.00 |
|  |  | 16-17 | 57,838 | <11 | 2,320,008 | 1.43 | 43,127 | <11 | 1,735,767 | 2.09 | 50,620 | <11 | 1,954,814 | 1.94 |
| Myocarditis/Pericarditis (1-21 day risk window, all settings) | Primary Series | 5-11 | 603,585 | <11 | 12,485,896 | 3.44 | 605,143 | <11 | 12,561,157 | 3.03 | 572,742 | <11 | 11,698,334 | 4.35 |
|  |  | 12-15 | 573,445 | 31 | 11,941,308 | **10.62*** | 653,742 | 23 | 13,640,542 | **9.06*** | 565,967 | 35 | 11,700,690 | **10.19*** |
|  |  | 16-17 | 291,721 | 28 | 6,071,623 | **12.65*** | 335,160 | 23 | 6,990,316 | **7.44*** | 286,151 | 13 | 5,912,687 | **3.47*** |
|  | Dose 1 | 5-11 | 318,545 | <11 | 6,590,589 | 3.25 | 335,584 | <11 | 6,969,330 | 1.80 | 310,676 | <11 | 6,341,416 | 3.20 |
|  |  | 12-15 | 302,195 | <11 | 6,282,142 | 2.60 | 356,836 | <11 | 7,433,347 | 2.89 | 302,684 | <11 | 6,254,311 | **4.90*** |
|  |  | 16-17 | 155,443 | <11 | 3,228,595 | **5.10*** | 187,217 | <11 | 3,897,594 | 4.07 | 155,846 | <11 | 3,217,800 | 0.98 |
|  | Dose 2 | 5-11 | 285,040 | <11 | 5,895,307 | 3.67 | 269,559 | <11 | 5,591,827 | 4.59 | 262,066 | <11 | 5,356,918 | 5.72 |
|  |  | 12-15 | 271,250 | <11 | 5,659,166 | **19.53*** | 296,906 | <11 | 6,207,195 | **16.45*** | 263,283 | <11 | 5,446,379 | **16.26*** |
|  |  | 16-17 | 136,278 | <11 | 2,843,028 | **21.21*** | 147,943 | <11 | 3,092,722 | **11.68*** | 130,305 | <11 | 2,694,887 | **6.45*** |
|  | Dose 3 | 5-11 | 11,443 | 0 | 94,226 | 0.00 | 1,341 | 0 | 27,092 | 0.00 | 19,047 | 0 | 249,882 | 0.00 |
|  |  | 12-15 | 86,157 | <11 | 1,753,863 | 2.52 | 63,977 | <11 | 1,320,089 | 14.01 | 79,609 | <11 | 1,588,091 | 4.47 |
|  |  | 16-17 | 57,844 | <11 | 1,180,160 | **9.93*** | 43,134 | <11 | 885,705 | 5.63 | 50,621 | <11 | 1,007,613 | 4.90 |
| Myocarditis/Pericarditis (1-7 day risk window, all settings) | Primary Series | 5-11 | 603,585 | <11 | 4,201,903 | 5.10 | 605,143 | <11 | 4,218,557 | 8.94 | 572,742 | <11 | 3,965,884 | 10.26 |
|  |  | 12-15 | 573,445 | 22 | 4,005,827 | **22.44*** | 653,742 | 14 | 4,569,263 | **16.43*** | 565,967 | 26 | 3,943,897 | **22.44*** |
|  |  | 16-17 | 291,721 | 22 | 2,037,440 | **29.60*** | 335,160 | 19 | 2,341,915 | **18.31*** | 286,151 | 11 | 1,993,951 | **8.72*** |
|  | Dose 1 | 5-11 | 318,545 | <11 | 2,218,337 | 4.81 | 335,584 | <11 | 2,340,724 | 5.33 | 310,676 | <11 | 2,150,922 | 4.72 |
|  |  | 12-15 | 302,195 | <11 | 2,110,944 | 1.93 | 356,836 | 0 | 2,494,047 | 0.00 | 302,684 | <11 | 2,109,334 | 6.45 |
|  |  | 16-17 | 155,443 | <11 | 1,085,398 | 7.58 | 187,217 | <11 | 1,307,806 | 6.91 | 155,846 | <11 | 1,085,979 | 2.91 |
|  | Dose 2 | 5-11 | 285,040 | <11 | 1,983,566 | 5.43 | 269,559 | <11 | 1,877,833 | 13.54 | 262,066 | <11 | 1,814,962 | 16.85 |
|  |  | 12-15 | 271,250 | <11 | 1,894,883 | **45.33*** | 296,906 | 14 | 2,075,216 | **36.17*** | 263,283 | <11 | 1,834,563 | **40.83*** |
|  |  | 16-17 | 136,278 | <11 | 952,042 | **54.68*** | 147,943 | <11 | 1,034,109 | **32.71*** | 130,305 | <11 | 907,972 | **15.66*** |
|  | Dose 3 | 5-11 | 11,443 | 0 | 48,031 | 0.00 | 1,341 | 0 | 9,287 | 0.00 | 19,047 | 0 | 107,282 | 0.00 |
|  |  | 12-15 | 86,157 | <11 | 594,686 | 7.41 | 63,977 | <11 | 445,495 | **40.59*** | 79,609 | <11 | 544,542 | 13.05 |
|  |  | 16-17 | 57,844 | <11 | 400,044 | **29.22*** | 43,134 | <11 | 299,708 | 8.20 | 50,621 | <11 | 347,047 | 14.22 |
| Myocarditis/Pericarditis (1-21 day risk window, IP, OP-ED) | Primary Series | 5-11 | 603,627 | 0 | 12,415,336 | 0.00 | 605,187 | <11 | 12,519,919 | 16.62 | 572,773 | <11 | 11,698,984 | 3.49 |
|  |  | 12-15 | 573,501 | 19 | 11,924,350 | **22.98*** | 653,800 | 12 | 13,627,693 | **16.86*** | 566,007 | 21 | 11,701,527 | **18.64*** |
|  |  | 16-17 | 291,767 | 22 | 6,063,130 | **20.76*** | 335,210 | 15 | 6,983,839 | **9.07*** | 286,174 | <11 | 5,913,153 | **4.98*** |
|  | Dose 1 | 5-11 | 318,568 | 0 | 6,554,190 | 0.00 | 335,609 | <11 | 6,950,073 | 14.71 | 310,693 | 0 | 6,341,772 | 0.00 |
|  |  | 12-15 | 302,223 | 0 | 6,272,569 | 0.00 | 356,870 | 0 | 7,425,502 | 0.00 | 302,703 | <11 | 6,254,710 | 4.98 |
|  |  | 16-17 | 155,466 | <11 | 3,223,290 | **8.87*** | 187,249 | <11 | 3,893,432 | 4.35 | 155,858 | <11 | 3,218,052 | 2.61 |
|  | Dose 2 | 5-11 | 285,059 | 0 | 5,861,146 | 0.00 | 269,578 | <11 | 5,569,846 | 19.10 | 262,080 | <11 | 5,357,212 | 7.62 |
|  |  | 12-15 | 271,278 | 19 | 5,651,781 | **48.61*** | 296,930 | 12 | 6,202,191 | **37.07*** | 263,304 | <11 | 5,446,817 | **34.34*** |
|  |  | 16-17 | 136,301 | <11 | 2,839,840 | **34.24*** | 147,961 | <11 | 3,090,407 | **15.01*** | 130,316 | <11 | 2,695,101 | **7.81*** |
|  | Dose 3 | 5-11 | 11,446 | 0 | 68,786 | 0.00 | 1,341 | 0 | 26,861 | 0.00 | 19,048 | 0 | 249,884 | 0.00 |
|  |  | 12-15 | 86,167 | <11 | 1,714,547 | 9.72 | 63,982 | <11 | 1,305,505 | 35.96 | 79,616 | 0 | 1,588,218 | 0.00 |
|  |  | 16-17 | 57,854 | <11 | 1,166,224 | 16.47 | 43,143 | <11 | 879,636 | 10.98 | 50,629 | <11 | 1,007,781 | 4.33 |
| Myocarditis/Pericarditis (1-7 day risk window, IP, OP-ED) | Primary Series | 5-11 | 603,627 | 0 | 4,176,754 | 0.00 | 605,187 | <11 | 4,207,157 | 48.87 | 572,773 | <11 | 3,966,101 | 10.29 |
|  |  | 12-15 | 573,501 | 19 | 3,999,983 | **68.36*** | 653,800 | <11 | 4,565,641 | **41.78*** | 566,007 | 19 | 3,944,177 | **50.01*** |
|  |  | 16-17 | 291,767 | 18 | 2,034,590 | **50.55*** | 335,210 | 13 | 2,340,241 | **23.39*** | 286,174 | <11 | 1,994,109 | **14.77*** |
|  | Dose 1 | 5-11 | 318,568 | 0 | 2,205,071 | 0.00 | 335,609 | <11 | 2,335,146 | 43.40 | 310,693 | 0 | 2,151,041 | 0.00 |
|  |  | 12-15 | 302,223 | 0 | 2,107,581 | 0.00 | 356,870 | 0 | 2,491,770 | 0.00 | 302,703 | <11 | 2,109,467 | 9.84 |
|  |  | 16-17 | 155,466 | <11 | 1,083,649 | 15.82 | 187,249 | <11 | 1,306,721 | 6.45 | 155,858 | <11 | 1,086,063 | 7.75 |
|  | Dose 2 | 5-11 | 285,059 | 0 | 1,971,683 | 0.00 | 269,578 | <11 | 1,872,011 | 55.92 | 262,080 | <11 | 1,815,060 | 22.49 |
|  |  | 12-15 | 271,278 | 19 | 1,892,402 | **144.90*** | 296,930 | <11 | 2,073,871 | **92.06*** | 263,304 | <11 | 1,834,710 | **96.22*** |
|  |  | 16-17 | 136,301 | <11 | 950,941 | **90.11*** | 147,961 | <11 | 1,033,520 | **44.75*** | 130,316 | <11 | 908,046 | **23.17*** |
|  | Dose 3 | 5-11 | 11,446 | 0 | 23,687 | 0.00 | 1,341 | 0 | 9,233 | 0.00 | 19,048 | 0 | 107,284 | 0.00 |
|  |  | 12-15 | 86,167 | <11 | 579,688 | 28.38 | 63,982 | <11 | 441,307 | 102.61 | 79,616 | 0 | 544,585 | 0.00 |
|  |  | 16-17 | 57,854 | <11 | 394,604 | **48.16*** | 43,143 | <11 | 297,977 | 31.73 | 50,629 | <11 | 347,103 | 12.56 |
| Narcolepsy | Primary Series | 5-11 | 603,613 | <11 | 19,492,848 | 3.00 | 605,180 | <11 | 19,963,122 | 0.47 | 572,762 | 0 | 18,207,458 | 0.00 |
|  |  | 12-15 | 573,450 | <11 | 18,629,851 | 0.50 | 653,703 | <11 | 21,635,586 | 0.73 | 565,952 | <11 | 18,280,657 | 0.76 |
|  |  | 16-17 | 291,655 | <11 | 9,533,109 | 0.82 | 335,066 | <11 | 11,254,465 | 0.76 | 286,088 | <11 | 9,348,810 | 0.60 |
|  | Dose 1 | 5-11 | 318,562 | <11 | 7,962,580 | 1.82 | 335,604 | <11 | 8,995,962 | 1.02 | 310,686 | 0 | 7,873,597 | 0.00 |
|  |  | 12-15 | 302,195 | <11 | 7,406,603 | 0.94 | 356,822 | <11 | 9,297,814 | 1.41 | 302,671 | <11 | 7,541,252 | 0.31 |
|  |  | 16-17 | 155,400 | <11 | 3,888,553 | 0.67 | 187,163 | <11 | 5,102,168 | 0.48 | 155,815 | <11 | 4,033,705 | 0.46 |
|  | Dose 2 | 5-11 | 285,051 | <11 | 11,530,268 | 3.82 | 269,576 | 0 | 10,967,160 | 0.00 | 262,076 | 0 | 10,333,861 | 0.00 |
|  |  | 12-15 | 271,255 | <11 | 11,223,248 | 0.21 | 296,881 | <11 | 12,337,772 | 0.21 | 263,281 | <11 | 10,739,405 | 1.08 |
|  |  | 16-17 | 136,255 | <11 | 5,644,556 | 1.15 | 147,903 | <11 | 6,152,297 | 0.99 | 130,273 | <11 | 5,315,105 | 0.70 |
|  | Dose 3 | 5-11 | 11,446 | 0 | 150,873 | 0.00 | 1,341 | 0 | 47,553 | 0.00 | 19,046 | 0 | 369,915 | 0.00 |
|  |  | 12-15 | 86,162 | <11 | 3,431,411 | 1.46 | 63,976 | 0 | 2,573,404 | 0.00 | 79,609 | <11 | 3,077,909 | 0.78 |
|  |  | 16-17 | 57,831 | <11 | 2,319,748 | 1.77 | 43,124 | <11 | 1,735,642 | 3.82 | 50,607 | <11 | 1,954,338 | 0.48 |
| Non-hemorrhagic stroke | Primary Series | 5-11 | 603,620 | 0 | 14,973,614 | 0.00 | 605,190 | <11 | 15,182,777 | 2.44 | 572,772 | 0 | 14,156,125 | 0.00 |
|  |  | 12-15 | 573,510 | 0 | 14,304,561 | 0.00 | 653,814 | 0 | 16,468,867 | 0.00 | 566,004 | 0 | 14,069,866 | 0.00 |
|  |  | 16-17 | 291,775 | <11 | 7,298,163 | 1.65 | 335,217 | 0 | 8,502,438 | 0.00 | 286,197 | <11 | 7,151,916 | 0.99 |
|  | Dose 1 | 5-11 | 318,565 | 0 | 7,238,863 | 0.00 | 335,611 | <11 | 7,844,822 | 4.61 | 310,691 | 0 | 7,096,122 | 0.00 |
|  |  | 12-15 | 302,226 | 0 | 6,798,090 | 0.00 | 356,878 | 0 | 8,227,890 | 0.00 | 302,701 | 0 | 6,848,826 | 0.00 |
|  |  | 16-17 | 155,470 | <11 | 3,524,039 | 3.42 | 187,253 | 0 | 4,394,094 | 0.00 | 155,873 | 0 | 3,578,081 | 0.00 |
|  | Dose 2 | 5-11 | 285,055 | 0 | 7,734,751 | 0.00 | 269,579 | 0 | 7,337,955 | 0.00 | 262,081 | 0 | 7,060,003 | 0.00 |
|  |  | 12-15 | 271,284 | 0 | 7,506,471 | 0.00 | 296,936 | 0 | 8,240,977 | 0.00 | 263,303 | 0 | 7,221,040 | 0.00 |
|  |  | 16-17 | 136,305 | 0 | 3,774,124 | 0.00 | 147,964 | 0 | 4,108,344 | 0.00 | 130,324 | <11 | 3,573,835 | 1.98 |
|  | Dose 3 | 5-11 | 11,446 | 0 | 87,634 | 0.00 | 1,341 | 0 | 33,429 | 0.00 | 19,048 | 0 | 291,412 | 0.00 |
|  |  | 12-15 | 86,169 | 0 | 2,248,357 | 0.00 | 63,983 | 0 | 1,697,487 | 0.00 | 79,623 | 0 | 2,092,148 | 0.00 |
|  |  | 16-17 | 57,855 | <11 | 1,536,784 | 8.80 | 43,144 | <11 | 1,152,674 | 21.01 | 50,632 | 0 | 1,327,126 | 0.00 |
| Pulmonary embolism (PE) | Primary Series | 5-11 | 603,628 | 0 | 15,109,619 | 0.00 | 605,187 | 0 | 15,312,029 | 0.00 | 572,776 | 0 | 14,156,230 | 0.00 |
|  |  | 12-15 | 573,504 | <11 | 14,338,531 | 1.06 | 653,808 | 0 | 16,512,834 | 0.00 | 566,002 | <11 | 14,069,828 | 0.56 |
|  |  | 16-17 | 291,724 | <11 | 7,314,350 | 1.58 | 335,190 | <11 | 8,524,694 | 1.79 | 286,159 | 0 | 7,150,951 | 0.00 |
|  | Dose 1 | 5-11 | 318,569 | 0 | 7,303,427 | 0.00 | 335,608 | 0 | 7,897,662 | 0.00 | 310,694 | 0 | 7,096,199 | 0.00 |
|  |  | 12-15 | 302,224 | 0 | 6,816,484 | 0.00 | 356,874 | 0 | 8,252,689 | 0.00 | 302,699 | 0 | 6,848,786 | 0.00 |
|  |  | 16-17 | 155,442 | <11 | 3,533,778 | 2.18 | 187,238 | <11 | 4,407,873 | 2.61 | 155,851 | 0 | 3,577,578 | 0.00 |
|  | Dose 2 | 5-11 | 285,059 | 0 | 7,806,192 | 0.00 | 269,579 | 0 | 7,414,367 | 0.00 | 262,082 | 0 | 7,060,031 | 0.00 |
|  |  | 12-15 | 271,280 | <11 | 7,522,047 | 2.02 | 296,934 | 0 | 8,260,145 | 0.00 | 263,303 | <11 | 7,221,042 | 1.10 |
|  |  | 16-17 | 136,282 | <11 | 3,780,572 | 1.02 | 147,952 | <11 | 4,116,821 | 0.93 | 130,308 | 0 | 3,573,373 | 0.00 |
|  | Dose 3 | 5-11 | 11,446 | 0 | 113,843 | 0.00 | 1,341 | 0 | 34,377 | 0.00 | 19,048 | 0 | 291,412 | 0.00 |
|  |  | 12-15 | 86,168 | 0 | 2,319,678 | 0.00 | 63,983 | 0 | 1,746,941 | 0.00 | 79,622 | 0 | 2,092,120 | 0.00 |
|  |  | 16-17 | 57,847 | <11 | 1,563,040 | 2.60 | 43,141 | 0 | 1,173,213 | 0.00 | 50,627 | 0 | 1,326,986 | 0.00 |
| Seizures/convulsions | Primary Series | 5-11 | 732,503 | 15 | 5,804,883 | 1.81 | 721,478 | 24 | 5,748,418 | 1.79 | 676,710 | 12 | 5,356,934 | 1.33 |
|  |  | 12-15 | 685,589 | 17 | 5,467,492 | 1.60 | 768,292 | 20 | 6,136,727 | 1.11 | 666,728 | 13 | 5,310,571 | 1.16 |
|  |  | 16-17 | 347,431 | 11 | 2,770,390 | 1.63 | 391,217 | 17 | 3,123,757 | 1.27 | 335,928 | <11 | 2,675,382 | 0.91 |
|  | Dose 1 | 5-11 | 388,213 | <11 | 3,076,139 | 2.26 | 401,399 | <11 | 3,199,876 | 1.98 | 369,270 | <11 | 2,922,238 | 1.22 |
|  |  | 12-15 | 363,843 | <11 | 2,900,710 | 1.42 | 422,576 | <11 | 3,375,193 | 0.91 | 358,999 | <11 | 2,859,080 | 1.33 |
|  |  | 16-17 | 186,243 | <11 | 1,484,375 | 2.22 | 220,499 | <11 | 1,760,130 | 1.33 | 184,243 | <11 | 1,467,105 | 1.04 |
|  | Dose 2 | 5-11 | 344,290 | <11 | 2,728,744 | 1.29 | 320,079 | <11 | 2,548,542 | 1.53 | 307,440 | <11 | 2,434,696 | 1.46 |
|  |  | 12-15 | 321,746 | <11 | 2,566,782 | 1.81 | 345,716 | <11 | 2,761,534 | 1.36 | 307,729 | <11 | 2,451,491 | 0.97 |
|  |  | 16-17 | 161,188 | <11 | 1,286,015 | 1.28 | 170,718 | <11 | 1,363,627 | 1.20 | 151,685 | <11 | 1,208,277 | 0.76 |
|  | Dose 3 | 5-11 | 13,572 | 0 | 34,157 | 0.00 | 1,476 | 0 | 11,690 | 0.00 | 21,413 | 0 | 141,013 | 0.00 |
|  |  | 12-15 | 96,123 | <11 | 747,307 | 3.04 | 66,287 | <11 | 527,774 | 0.73 | 85,908 | <11 | 672,632 | 1.44 |
|  |  | 16-17 | 65,777 | <11 | 516,066 | 0.86 | 45,550 | 0 | 361,811 | 0.00 | 55,387 | 0 | 434,725 | 0.00 |

Abbreviation: RR, rate ratio; IP, inpatient; OP-ED, Outpatient Emergency Department

*Data cuts: CVS Health data through 5/31/2022, HealthCore data through 5/6/2022, and Optum data through 6/25/2022*

*indicates the AESI has signaled in the data.

Cell sizes 1-10 were masked for confidentiality

**Supplemental Table 5. Descriptive Outcome Counts, Overall and by Data Partners**

| **Outcome** | **All Data Partners** | | **CVS (data through 5/31/22)** | | **HealthCore (data through 5/6/22)** | | **Optum (data through 6/25/22)** | |
| --- | --- | --- | --- | --- | --- | --- | --- | --- |
|  | **# of Vaccine Doses** | **# of Vaccine Doses** | **# of Vaccine Doses** | **# of Vaccine Doses** | **# of Vaccine Doses** | **# of Outcomes** | **# of Vaccine Doses** | **# of Outcomes** |
| **AMI** | 4,904,152 | 1,625,585 | 1,703,210 | 1,575,357 | 1,575,357 | <11 | 1,575,357 | 0 |
| **GBS** | 4,904,135 | 1,625,574 | 1,703,207 | 1,575,354 | 1,575,354 | <11 | 1,575,354 | <11 |
| **Hemorrhagic Stroke** | 4,904,039 | 1,625,548 | 1,703,163 | 1,575,328 | 1,575,328 | <11 | 1,575,328 | <11 |
| **Kawasaki Disease** | 4,903,075 | 1,625,217 | 1,702,843 | 1,575,015 | 1,575,015 | 19 | 1,575,015 | 24 |
| **MIS-C** | 4,903,902 | 1,625,518 | 1,703,107 | 1,575,277 | 1,575,277 | <11 | 1,575,277 | <11 |
| **Transverse Myelitis** | 4,904,143 | 1,625,584 | 1,703,206 | 1,575,353 | 1,575,353 | 0 | 1,575,353 | <11 |
| **Unusual Site Thromboses** | 4,904,140 | 1,625,582 | 1,703,209 | 1,575,349 | 1,575,349 | 0 | 1,575,349 | 0 |

Cell sizes 1-10 were masked for confidentiality
